# Low levels of circulating IgG against bacterial GAPDH and elevated IL-10 levels are associated with increased susceptibility to *Escherichia coli* bacteraemia

**DOI:** 10.64898/2025.12.15.25342302

**Authors:** Ana Fidalgo, Filipa Lemos, Joana B. Nunes, Carla Teixeira, Cristiana Nogueira, Hugo Osório, Sofia Botelho Moniz, Cristina Isabel Silva, Carolina Lemos, Pedro Castanheira, Marta Vieira, Pedro Madureira

## Abstract

**Background:** While antimicrobial resistance is an increasingly urgent problem, with *Escherichia coli* infections representing a major priority, the development of effective vaccines has proven both challenging and largely unsuccessful. Given accumulating evidence supporting the immunosuppressive role of extracellular bacterial glyceraldehyde-3-phosphate dehydrogenase (GAPDH), we aimed to characterize individual susceptibility to *E. coli* infections based on the presence of naturally induced antibodies against this protein.

**Methods:** We conducted an observational case-control study including 62 individuals with *E. coli* bacteraemia (cases) and 124 age- and sex-matched controls without infection. Detection of GAPDH was performed in plasma samples, and plasma interleukin (IL)-10 levels and anti-GAPDH IgG (titers and concentrations) were quantified. Associations between anti-GAPDH IgG levels and infection were evaluated using logistic regression, and individual disease risk was estimated with a multivariate model incorporating IL-10 detection and low levels of anti-GAPDH IgG.

**Results:** *E. coli* GAPDH was detected in the plasma of cases from which purified colonies of *E. coli* were isolated. IL-10 levels were significantly higher (p < 0.0001) in cases while anti-GAPDH IgG levels were significantly lower (p < 0.0001) comparing with controls. Logistic regression analysis revealed a strong inverse association between anti-GAPDH IgG and the diagnosis of *E. coli* bacteraemia (adjusted OR = 0.18, 95% CI 0.08-0.37) and a protective threshold of 1.0 μg/mL was estimated, below which, 95.2% of cases were classified. In a multivariate logistic regression model, detection of IL-10 was strongly associated with infection risk (OR = 629, 95% CI 139–5355), while low anti-GAPDH IgG showed a trend towards increased risk (OR = 4.26, 95% CI 0.91–30.6). This model allowed the estimation of individual probabilities and absolute risk of *E. coli* bacteraemia using a combining biomarker based on levels of anti-GAPDH IgG and IL-10.

**Conclusions:** This study provides the first evidence in humans of the protective potential of circulating anti-GAPDH antibodies, which supports GAPDH as a promising target for an alternative vaccination strategy to prevent *E. coli* infections.

## BACKGROUND

Antimicrobial resistance (AMR) is a major global public health concern, associated with high morbidity and mortality, and with an increasingly large financial impact (1). Among the most life-threatening pathogens, *Escherichia coli* is a leading cause of both community- and hospital-acquired infections, including urinary tract infections, bloodstream infections, and ventilator-associated pneumonia (2). These infections are highly prevalent and increasingly resistant to available treatments, with limited therapeutic options and no effective preventive measures. For these reasons, since 2017 and further confirmed in the latest 2024 report, the World Health Organization has classified *E. coli* as a critical pathogen for the development of novel therapeutic and preventive strategies (3,4).

Vaccination is a key strategy to prevent infections and has a significant potential to reduce AMR, by reducing antibiotic use and, consequently, the selective pressure for resistant strains. Additionally, novel vaccines can be the solution to infections caused by pathogens for which no effective treatments are currently available (5,6), with a potential to prevent up to 10% of the estimated 4.95 million annual deaths associated with AMR (5).

However, despite extensive efforts, no licensed vaccine is currently available to prevent *E. coli* infections (6–8). O-antigen-based vaccines have been extensively investigated and advanced to phase 3 with the multivalent ExPEC9V, which targets the serotypes most frequently associated with urinary tract infections (UTI) and bacteraemia (9). Although immunogenic and well tolerated, this vaccine failed to be efficient in preventing invasive *E. coli* disease in phase 3 of clinical trials (10). A more recent O-antigen-based formulation, the ExPEC10V vaccine, has completed phase 1/2a (NCT03819049) demonstrating robust immunogenicity and acceptable safety profile trials (11). The SEQ-400 vaccine, which targets another surface antigen, the FimH adhesin, is also currently under clinical evaluation and has demonstrated favourable safety and immunogenicity in phase 1 (12).

Different serological studies have reported that infections caused by *E. coli* and other *Enterobacteriaceae* promote an increase in serum antibodies against surface antigens. However, these antibodies do not correlate with protection against recurrent infections caused by the same pathogens (13–22). Together with the lack of efficacy of immunogenic vaccine candidates, these findings reinforce that antibody responses against bacterial surface antigens are not protective *per se*.

Glyceraldehyde-3-phosphate dehydrogenase (GAPDH) is a moonlighting protein that has essential metabolic functions but is also secreted by pathogenic strains, with important virulence functions (23,24) associated with an overall immunosuppression that is essential for host colonization and evasion (25). Extracellular bacterial GAPDH has been emerging as a promising vaccine candidate for two main reasons: *i)* it is a well conserved protein excreted by different bacteria (24,26), which overcomes the problem of serotype specificity and diversity and *ii) in vivo* murine studies show that anti-GAPDH antibodies counteract early immunosuppression induced by interleukin (IL)-10 upon infection and increase the survival capacity to infection (25,27,28)

As increasing evidence supports the virulent role of extracellular GAPDH in the context of infections caused by *E. coli* (25, unpublished observations), we conducted an observational study to characterize the association between circulating antibodies against bacterial GAPDH and individual susceptibility to systemic *E. coli* infections.

## METHODS

### Study design, Participants and Samples

A retrospective matched case-control observational study was conducted comprising the analysis of peripheral blood from a total of 186 individuals aged > 30 years, defined as cases or controls. Individuals diagnosed with bacteraemia, confirmed with a positive blood culture, were considered cases. Individuals without a diagnosis of infection and with negative blood cultures were considered controls.

Samples and the respective anonymized clinical information were obtained from Discovery Life Sciences (Huntsville, AL, USA) (n=78) and Unilabs (Porto, Portugal) (n=108). Peripheral blood was centrifuged at 13,000 rpm for 10 minutes at 4 °C. Plasma was separated, 0.2 μm filtered and stored at –80 °C until analysis.

Cases were stratified by age groups in ten-year intervals (from 30 to 100 years) and each case was age matched with two controls from the same age group, and ensuring an equal distribution of individuals by sex within the two groups.

### Production and purification of recombinant GAPDH

Recombinant *E. coli* GAPDH (rGAPDH) was expressed in *E. coli* BL21 Star™ (DE3) (ThermoFisher Scientific) using a pET28a expression plasmid containing the codon-optimized *gapC* gene sequence fused to a C-terminal hexahistidine tag. Transformed bacteria were grown in Luria–Bertani (LB) Miller broth supplemented with 50 µg/mL kanamycin at 37 °C with shaking until reaching an optical density of 0.5 at 600 nm. Protein expression was induced after lowering the temperature to 18 °C and by adding IPTG to a final concentration of 0.1 mM, and the culture was grown overnight at 18 °C under constant shaking.

Cells were collected by centrifugation (6,000 × g, 20 min) and resuspended in binding buffer (20 mM sodium phosphate, pH 7.4, 500 mM NaCl, 15 mM imidazole). Lysozyme was added before storage at −20 °C. After thawing, MgCl₂ and DNase I were added to promote DNA degradation. The lysate was clarified by ultracentrifugation (50,000 × g) and filtered (0.22 µm) prior to purification.

Purification of rGAPDH was performed using a HisTrap™ HP affinity column (Cytiva) on an ÄKTA Pure 25 system. Bound protein was eluted using a stepwise imidazole gradient, with rGAPDH eluting at 150 mM imidazole step. A polishing purification step was carried out by size-exclusion chromatography (HiLoad Superdex 200 26/600 pg, Cytiva) using PBS as the mobile phase. Protein purity was confirmed by SDS-PAGE.

### Production and purification of anti-rGAPDH IgG

To produce antibodies that recognize the extracellular form of *E. coli* GAPDH, a goat was immunized intramuscularly with 50 μg of *E. coli* rGAPDH, following the SICGEN protocol. The immunization consisted of four doses, with three-week intervals between doses. Antibody titers were assessed two weeks after the last dose.

Affinity purification of antibodies was performed on a CNBr Sepharose column (Cytiva, Upsala, Sweden) to which *E. coli* rGAPDH was covalently immobilized, using an ÄKTA Pure 25 system. The eluted fraction was desalted into binding buffer (20 mM phosphate, 300 mM NaCl, pH 7.4) using two tandem HiTrap Phast Desalting 5 mL columns (Cytiva). For IgG isolation, the desalted sample was loaded onto a HiTrap Protein G HP 1 mL column (Cytiva) and eluted with 100 mM glycine-HCl, pH 2.7 into tubes containing 1 M Tris-HCl, pH 9.0, to immediately neutralize the eluate. A final buffer exchange to 0.9% NaCl was performed using the same tandem desalting columns.

A portion of the purified antibodies was biotinylated using the EZ-Link NHS-PEG4 Biotinylation Kit (Thermo Fisher Scientific) according to the manufacturer’s instructions, for subsequent use in a streptavidin-based detection ELISA, described below.

Antibody concentration (both after purification and after biotinylation) was determined by measuring absorbance at 280 nm using a NanoDrop™ One Spectrophotometer (Thermo Fisher Scientific). Antibody concentration (both after purification and after biotinylation) was determined by measuring absorbance at 280 nm using a NanoDrop™ One Spectrophotometer (Thermo Fisher Scientific).

### Detection of extracellular GAPDH by mass spectrometry

Each sample was processed for proteomic analysis following the solid-phase-enhanced sample-preparation (SP3) protocol and enzymatically digested with trypsin/LysC as previously described (29).

Protein identification and quantitation was performed by nanoLC-MS/MS equipped with a Field Asymmetric Ion Mobility Spectrometry - FAIMS interface. This equipment is composed of a Vanquish Neo liquid chromatography system coupled to an Eclipse Tribrid Quadrupole, Orbitrap, Ion Trap mass spectrometer (Thermo Scientific, San Jose, CA). 250 nanograms of peptides of each sample were loaded onto a trapping cartridge (PepMap Neo C18, 300 μm x 5 mm i.d., 174500, Thermo Scientific, Bremen, Germany). Next, the trap column was switched in-line to an Aurora Frontier XT 60 cm, 75 μm (AUR4-60075C18-XT) chromatographic separation column. A 116 min separation was achieved by mixing A: 0.1% FA and B: 100% ACN, 0.1% FA with the following gradient at a flow of 250 nL/min: 2 min (0% B to 4% B), 20 min (4% B to 12% B), 65 min (12% B to 28% B), 11 min (28% B to 45% B), 2 min (45% B to 85 % B) and 16 min at 99% B. Subsequently, the column was equilibrated with 0% B. Data acquisition was controlled by Xcalibur 4.7 and Tune 4.2.4321 software (Thermo Scientific, Bremen, Germany).

MS results were obtained following a Data Dependent Acquisition - DDA procedure. MS acquisition was performed with the Orbitrap detector at 120,000 resolution in positive mode, quadrupole isolation, scan range (*m/z*) 375-1,500, RF Lens 30%, standard AGC target, maximum injection time was set to auto, 1 microscan, data type profile and without source fragmentation. FAIMS mode: standard resolution, total carrier gas flow: static 4L/min, FAIMS CV: −45, −60 and −75 (cycle time, 1 s). Internal Mass calibration: Run-Start Easy-IC. Filters: MIPS, monoisotopic peak determination: peptide, charge state: 2-7, dynamic exclusion 30s, intensity threshold, 5.0e^3^. MS/MS data acquisition parameters: quadrupole isolation window 1.8 (m/z), activation type: HCD (30% CE), detector: ion trap, IT scan rate: rapid, mass range: normal, scan range mode: auto, normalized AGC target 100%, maximum injection time: 35 ms, data type centroid.

The raw data was processed using the Proteome Discoverer 3.1.1.93 software (Thermo Scientific) and searched against the UniProt database for the *Homo sapiens* reviewed proteome (2024_01 with 20,418 entries) and the *E coli* O1K1 APEC proteome (2023_04 with 4,871 entries). A common protein contaminant list from MaxQuant was also included in the analysis. The MSPepSearch and Sequest HT and the search engines were used to identify tryptic peptides. The ion mass tolerance was 10 ppm for precursor ions and 0.5 Da for fragment ions. The maximum allowed missing cleavage sites was set to two. Cysteine carbamidomethylation was defined as constant modification. Methionine oxidation, deamidation of glutamine and asparagine, peptide terminus glutamine to pyroglutamate, and protein N-terminus acetylation, Met-loss, and Met-loss+acetyl were defined as variable modifications. Peptide confidence was set to high. The processing node Percolator was enabled with the following settings: maximum delta Cn 0.05; target FDR (strict) was set to 0.01 and target FDR (relaxed) was set to 0.05, validation based on *q*-value. Protein label-free quantitation was performed with the Minora feature detector node at the processing step. Precursor ions quantification was performed at the consensus step with the following parameters: inclusion of unique plus razor peptides, precursor abundance based on intensity, and normalization based on total peptide amount.

### Detection of extracellular GAPDH by ELISA

Ninety-six-well plates were coated with 5 µg/mL of goat anti-rGAPDH of *E. coli* IgG in PBS (pH 7.4) and incubated overnight (O/N) at 4 °C. Plates were then washed with 0.05% Tween-20 in PBS (PBST) for four wash cycles and blocked with 1% BSA in PBS for 1 hour at room temperature (RT). After another four wash cycles, human plasma samples diluted 1:2 in 1% BSA in PBS were added and incubated for 2 hours at RT. Plates were washed four times before adding 0.1 μg/mL of biotinylated goat anti-rGAPDH IgG in 1% BSA in PBS for 1 hour at RT. Following four wash cycles, streptavidin-HRP (Southern Biotech) diluted 1:5,000 in 1% BSA in PBS was added and incubated for 1 hour at RT, protected from light. A final washing consisting of five wash cycles was performed before adding TMB substrate. The reaction was stopped after 5 minutes with 1M of H_2_SO_4_ (1:1 volume ratio to TMB). Absorbance was measured at 450 nm using the Multiskan^TM^ FC Microplate Photometer (Thermo Scientific). The lower limit of detection (LLOD) was defined as 0.07.

### Quantification of IL-10

IL-10 quantification in human plasma was performed by ELISA (eBioscience), according to the manufacturer’s instructions. Optical density at 450 nm was measured in the Multiskan^TM^ FC Microplate Photometer (Thermo Scientific). Concentration of IL-10 was calculated by linear regression analysis, plotting absorbance (y-axis) against concentration (x-axis). The LLOD was 0.60 pg/mL and to the concentrations below it was given an arbitrary value of half the LLOD, 0.30 pg/mL.

### Quantification of anti-GAPDH human IgG

ELISA 96-well microplates were coated O/N at 4 °C with 5 µg/mL rGAPDH of *E. coli* in PBS (pH 7.4). Plates were washed with PBST for four wash cycles and blocked with 1% BSA in PBS for at least 1 hour at RT. After another four wash cycles, seven dilutions of human plasma (starting from a 1:10 dilution, followed by six 3-fold serial dilutions in 1 % BSA in PBS) were added and incubated for 2 hours at RT. Plates were washed with four wash cycles before adding goat anti-human IgG-HRP (Southern Biotech) diluted 1:5,000 in 1% BSA in PBS and incubating for 1 hour at RT. A final five-cycle PBST wash was performed before adding TMB substrate. The reaction was stopped after 5 minutes with 1 M of H_2_SO_4_ (1:1 volume ratio to TMB).

In parallel, a standard curve of human IgG (Southern Biotech) was included on all plates. Coating was performed with 5 μg/mL of goat Fab anti-human IgG (Southern Biotech), and seven dilutions of human IgG were added (starting at 0.1 µg/mL, followed by six 2-fold dilutions in 1% BSA in PBS), following the same procedure as described for plasma samples. Absorbance at 450 nm was measured in the Multiskan^TM^ FC Microplate Photometer (Thermo Scientific).

Titers of human anti-GAPDH IgG were defined as the highest dilution giving an absorbance above 0.09 at 450 nm. Concentration of human anti-GAPDH IgG was calculated by linear regression analysis, plotting absorbance (y-axis) against concentration (x-axis). The lower limit of quantification (LLOQ) was 0.003 μg/mL and to the concentrations below it was given an arbitrary value of half the LLOQ, 0.0015 μg/mL.

### Statistical analysis

Chi-square test was used to compare the distribution of participants according to sex between controls and cases. Mean age differences, concentration of IL-10, titers and concentration of IgG were evaluated using the unpaired t test with Welch’s correction. For IgG titers and IgG concentrations, values of individual concentrations were logarithmically transformed prior to analysis and back transformed for graphic representation. Fisher’s exact test was used to evaluate the association between detection of IL-10 and diagnosis of *E. coli* bacteraemia.

Associations between anti-GAPDH IgG concentration and diagnosis of bacteraemia were estimated by odds ratio (OR) and the corresponding 95% confidence intervals (CI), by simple logistic regression, and adjusted for age and sex, by multiple logistic regression. Receiver operating characteristic (ROC) curve and the area under de curve (AUC) were used to evaluate the predictive power of the model. Predicted probabilities of *E. coli* bacteraemia across the observed range of anti-GAPDH IgG concentrations were derived from these models, with age fixed at the median (70 years) and sex fixed at the most frequent category (woman) in the adjusted model.

Levels of anti-GAPDH IgG were categorised according to the distribution of this variable in the control group as follows: Q1, very low (< 0.22 μg/mL); Q2, low (0.22 - < 0.38 μg/mL), Q3, high (0.38 - < 0.72 μg/mL) and very high (≥ 0.72 μg/mL). A threshold of < 0.10 μg/mL was used as the reference group for estimation of OR and 95% CI.

The cumulative distribution of anti-GAPDH IgG concentrations was plotted only for cases, with relative frequencies grouped in bins of 0.20 μg/mL.

Association between detection of IL-10 and low levels of anti-GAPDH IgG was evaluated using the Chi-squared test. A multivariate logistic regression model combining IL-10 and low levels of anti-GAPDH IgG was fitted unadjusted and adjusted to age and sex. ROC curves and AUC were used to evaluate the performance of the models and calibration curves evaluated the model discrimination by plotting observed and predicted probabilities. Point-estimated predicted probabilities from the unadjusted multivariate model were converted into absolute risks calibrated to the population incidence of *E. coli* bacteraemia (48 cases per 100,000 person-year (30)). Uncertainty in regression coefficients was accounted by simulating 10,000 draws from the estimated multivariate normal distribution. Median predicted probabilities and absolute risks with 95% CI were calculated for each combination of biomarker profiles.

Two-sided p-values < 0.05 were considered statistically significant. Statistical analyses were performed using GraphPad Prism 10.6.0 and RStudio 2025.09+387 for macOS.

## RESULTS

### Participant characteristics

*E. coli* bacteraemia accounts for one-quarter of the diagnosed bacteraemia episodes, with an overall incidence rate of 48 cases per 100,000 person-years (30). Incidence increases with age, ranging from 110 to 319 cases per 100,000 person-years in individuals over 60 to those over 80. Overall incidence is higher in women, although this sex difference is not observed in individuals aged over 60 years (30).

In this observational study, 62 individuals diagnosed with *E. coli* bacteraemia were included and classified as cases. The mean ± SD age of cases was 72 ± 14 years (median 74 years, range 62-83 years) and 61.3% were women (Table 1). Considering the potential bias induced by age-related incidence, we matched each case with 2 controls by age, while ensuring that no significant differences exist in the proportion of men and women between groups. As a result, 124 individuals were selected as controls, with a mean ± SD age of 68 ± 12 years (median 70 years, range 61-77 years) and 63.7% were women (Table 1).

**Table 1.**
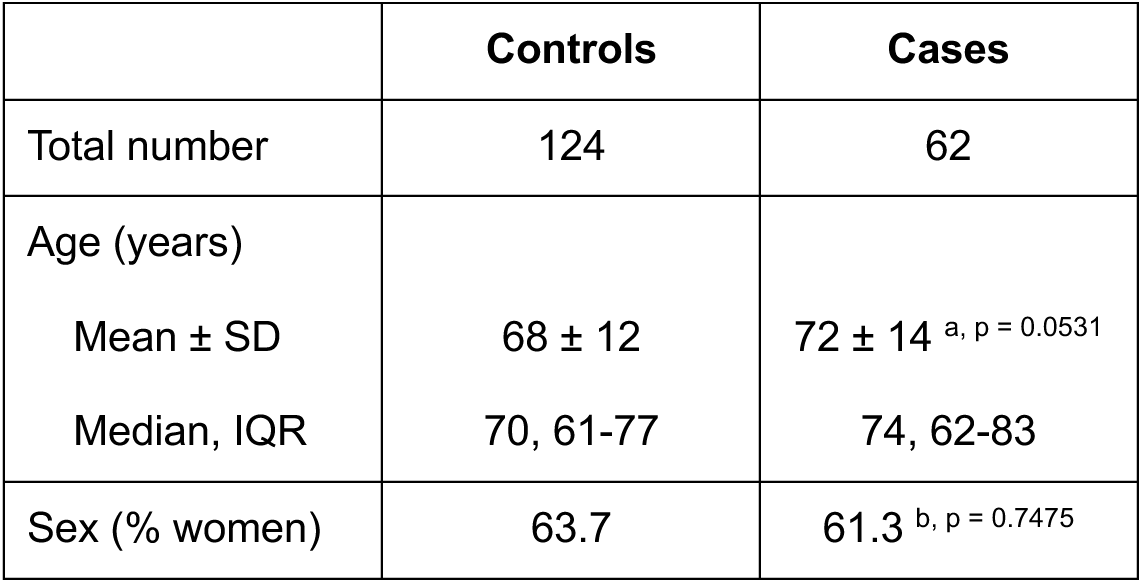
Demographic characteristics of controls and cases. Age is presented as mean ± standard deviation (SD) and median with interquartile range (IQR). Sex is presented as relative frequencies ( %). Mean differences compared with controls were tested using Welch’s t test^a^, and differences in proportions were tested using the Chi-squared test^b^.

### Detection of GAPDH in the blood of individuals with bacteraemia

Accumulating evidence suggests that various pathogens can produce and excrete an extracellular form of GAPDH that has important virulence functions (24,25,29, unpublished observations). Based on this, we hypothesize that circulating bacterial GAPDH should be detectable in individuals with *E. coli* bacteraemia.

To investigate this, whole blood samples from participants were plated on Todd-Hewitt agar to confirm the microbiological diagnosis. Pure *E. coli* colonies were isolated from seven cases, in which *E. coli* was identified as the sole pathogen in the sample. Therefore, we performed mass spectrometry analysis of these plasma samples to evaluate the presence of bacterial GAPDH. As shown in Table 2, circulating extracellular bacterial GAPDH was detected in the seven samples analysed.

**Table 2.** Summary of mass spectrometry results of the detection of extracellular bacterial GAPDH. GAPDH was detected in plasma samples from individuals diagnosed with *E. coli* bacteraemia. Columns include sequence coverage, number of peptide-spectrum matches (PSMs), number of unique peptides, annotated peptide sequences and PSMs corresponding to each peptide.

| Sample | Coverage [%] | PSMs | Number of unique peptides | Annotated sequences of peptides<br>(bold highlights peptides unique to *gapC*) | PSMs | Reference sequence |
| --- | --- | --- | --- | --- | --- | --- |
| S15EC | 25 | 9 | 6 | [K].VGINGFGR.[I] | 6 | B7MMQ3 |
|  |  |  |  | [K].HDSNYGPFWSVDYTEDSLIVDGK.[S] | 1 |  |
|  |  |  |  | [K].SQAHL DAGAK.[K] | 1 |  |
|  |  |  |  | [K].VTAE EVNNALK.[KQN] | 1 |  |
|  |  |  |  | [K].SI AVYAEK.[E] | 1 |  |
|  |  |  |  | [K].VLISAPAGEMK.[T] | 1 |  |
|  |  |  |  | [K].AIGLVIPELSGK.[L] | 6 |  |
| S70EC | 8 | 6 | 2 | [KT].VGINGFGR.[I] | 9 | B7MMQ3 |
|  |  |  |  | [K].SQAHL DAGAK.[K] | 1 |  |
|  |  |  |  | [K].SI AVYAEK.[E] | 1 |  |
| S71EC | 15 | 9 | 5 | [KT].VGINGFGR.[I] | 5 | B7MMQ3 |
|  |  |  |  | [K].VTAE EVNNALK.[KQN] | 1 |  |
|  |  |  |  | [K].SQAHL DAGAK.[K] | 1 |  |
|  |  |  |  | [K].SI AVYAEK.[E] | 1 |  |
|  |  |  |  | [K].AIGLVIPELSGK.[L] | 1 |  |
|  |  |  |  | [K].KVTAEEVNNALK.[KQN] | 1 |  |
|  |  |  |  | [K].VGINGFGR.[I] | 2 |  |
| S73EC | 2 | 1 | 0 | [KT].VGINGFGR.[I]* | 2 | B7MMQ3 |
| S74EC | 2 | 1 | 0 | [KT].VGINGFGR.[I]* | 2 | B7MMQ3 |
| S75EC | 16 | 5 | 3 | [KT].VGINGFGR.[I] | 2 | B7URG5 |
|  |  |  |  | [K].ILAYLLK.[H] | 1 |  |
|  |  |  |  | [K].TGSVTELVSILGK.[K] | 1 |  |
|  |  |  |  | [K].HDSNYGPFWSVDYTEDSLIVDGK.[S] | 1 |  |
|  |  |  |  | [K].VGINGFGR.[I] | 1 |  |
| S76EC | 5 | 3 | 1 | [KT].VGINGFGR.[I] | 4 | B7MMQ3 |
|  |  |  |  | [K].SI AVYAEK.[E] | 1 |  |

Detecting bacterial proteins at low concentrations, such as the expected levels of extracellular bacterial GAPDH, in complex samples like human blood, is particularly challenging. The complexity of the blood matrix can impact parameters that provide confidence in the detection of extracellular GAPDH, such as the percentage of coverage, and may also limit the identification of unique peptides corresponding to extracellular GAPDH. Conscious of these limitations and to increase the confidence in these findings, we also performed the detection of GAPDH by ELISA, using polyclonal IgG against bacterial GAPDH. Once again, GAPDH was detected in all samples (Figure 1), supporting the hypothesis that bacterial excretion of GAPDH represents a key step in its virulence mechanism, which also indicates its potential role as an early biomarker for infection.

**Figure 1.**
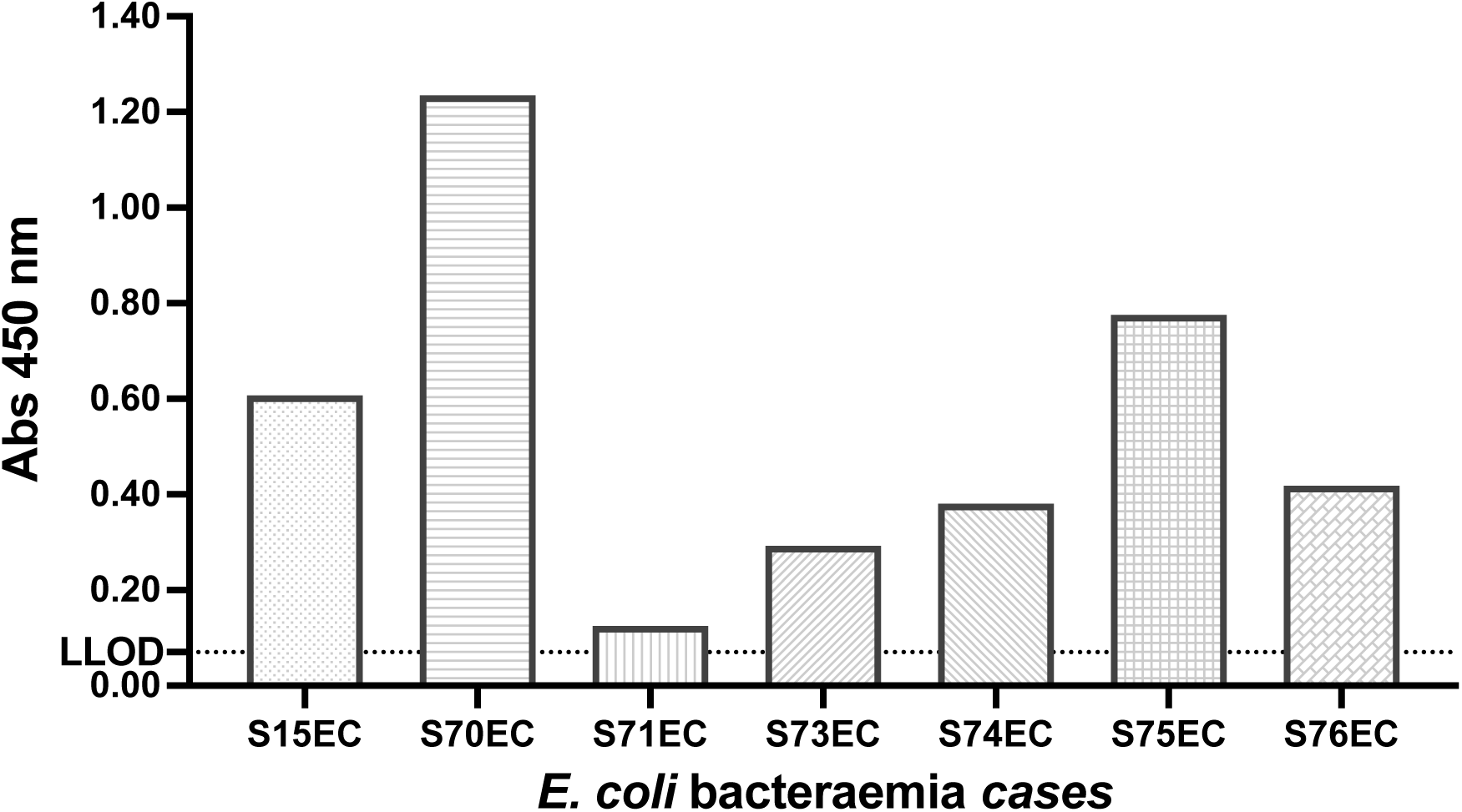
Detection of extracellular bacterial GAPDH in plasma from cases with pure *E. coli* isolates (n=7). Absorbance values at 450 nm correspond to ELISA readings using anti-GAPDH IgG purified from goat immunized with bacterial GAPDH. The dashed line indicates the lower limit of detection (LLOD) of the assay (0.07).

In addition to detecting extracellular bacterial GAPDH in the bloodstream of individuals with *E. coli* bacteraemia, we also evaluate the degree of conservation of this protein across different clinical isolates. To this end, we sequenced the *gapC* gene from each clinical isolate and performed a comparative analysis of the predicted amino acid sequences. Sequence alignments were conducted using the BLOSUM50 substitution matrix, revealing a high degree of similarity and identity among isolates (Supplementary data Table S1).

### High levels of circulating IL-10 are associated with the diagnosis of bacteraemia

Early and elevated production of IL-10 has been described in patients with severe infections and associated with higher mortality rate (32–37) and proposed as a candidate biomarker for the early diagnosis of bacterial infections (38).

In our study, IL-10 was only detected in 2 (1.63%) controls but detected in 56 (90.3%) of cases, which reveals a significant association between detectable concentrations of IL-10 at the time of blood collection and the diagnosis of *E. coli* bacteraemia (Fisher’s exact test p-value < 0.0001, Supplementary data Table S2). And, as expected and observed in Figure 2, the concentrations of IL-10 were significantly higher in cases comparing with controls (78.0 vs 0.92 pg/mL, p-value < 0.0001), which is in accordance with other studies and aligns with the hypothesis suggesting bacterial GAPDH as an immunosuppressive virulence factor (25,27,28).

**Figure 2.**
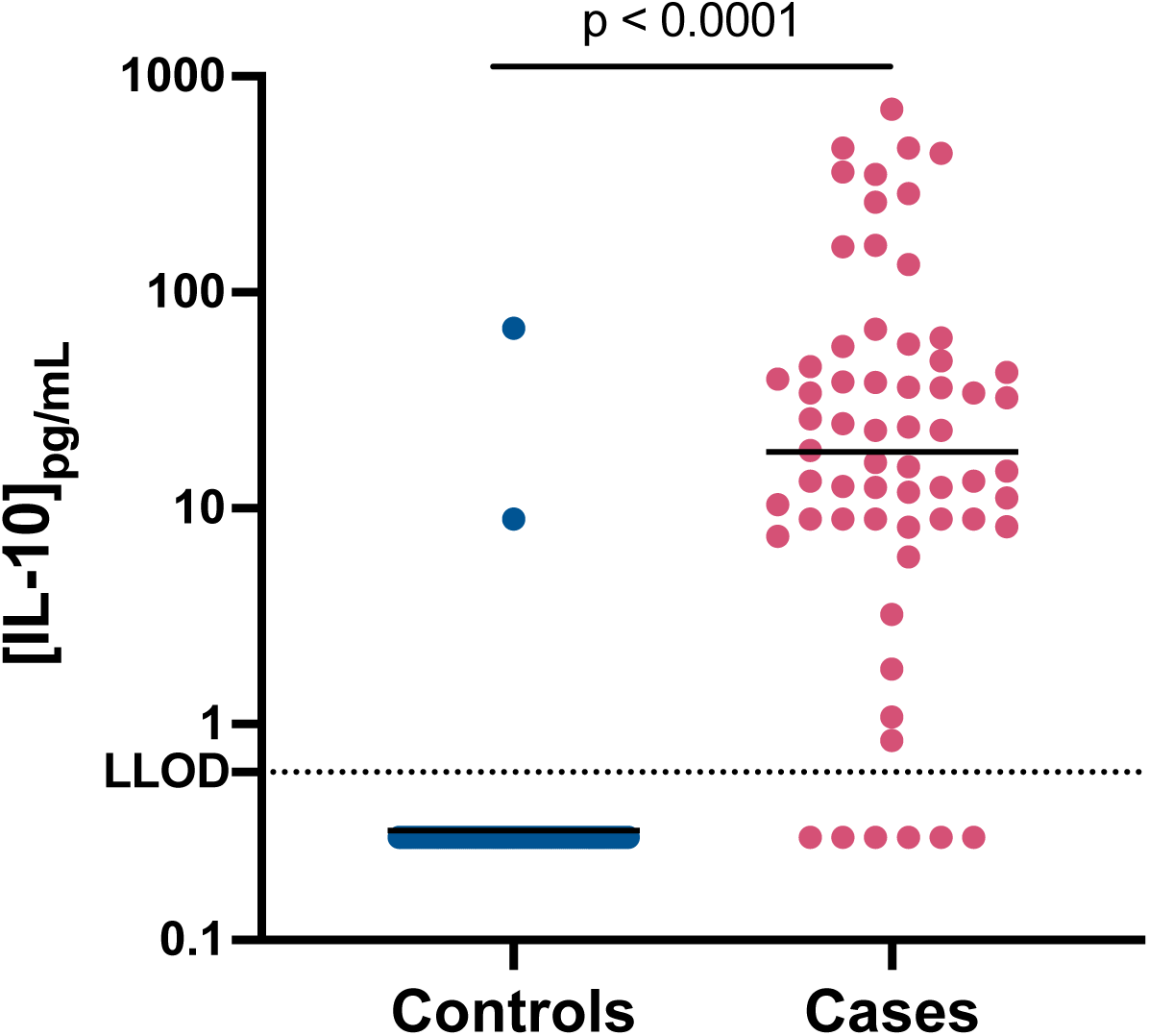
Detection of plasma IL-10 in controls and cases. Values are shown as individual concentrations with the horizontal bars representing the mean. Dashed line indicates the lower limit of detection (LLOD), 0.6 pg/mL; concentrations below the LLOD were assigned an arbitrary value of half the LLOD. Statistical analysis was performed using an unpaired t test with Welch’s correction.

### Individuals with bacteraemia have decreased concentrations of circulating anti-GAPDH IgG

To date, vaccines targeting *E. coli* surface antigens have proven immunogenic but ineffective, as antibody presence does not correlate with protection (9,10,13,14,16,18). In parallel, cumulative evidence supports our hypothesis that the immunosuppressive function of extracellular bacterial GAPDH impairs the immune system’s ability to mount an effective response (27,28), thereby limiting the protective potential of antibodies targeting structural epitopes.

In this study, we detected bacterial GAPDH in the plasma of individuals with confirmed *E. coli* infection. Also, infected individuals exhibited elevated IL-10 levels, which is consistent with the proposed GAPDH-induced immunosuppression (25,27,28). Our hypothesis suggests that the elevated IL-10 levels observed in infected individuals are driven by the absence of antibodies capable of neutralizing bacterial GAPDH, thereby allowing its immunosuppressive virulence function to persist. To address this, we quantified circulating anti-GAPDH IgG titers and concentrations, which revealed significant differences between controls and cases. Regarding the concentration, and as shown in Figure 3, a 2.7-fold reduction was observed in the geometric mean concentration (GMC) of anti-GAPDH IgG (0.38 vs 0.14 μg/mL, p-value < 0.0001) between controls and cases. And a very similar reduction (2.8-fold) was observed in the geometric mean titer (GMT) of anti-GAPDH IgG (228 vs 82, p-value < 0.0001) (Supplementary data Figure S1).

**Figure 3.**
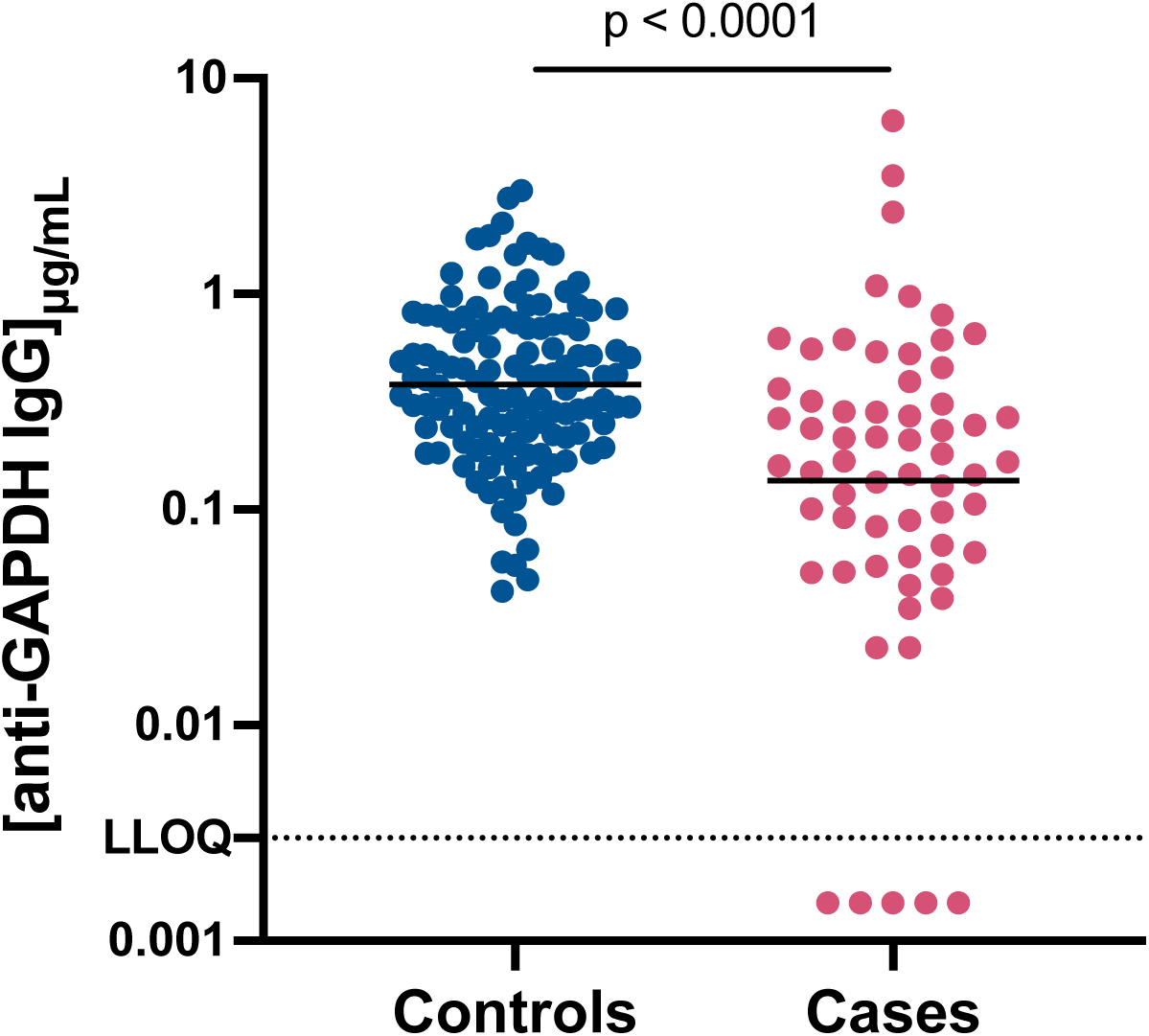
Circulating anti-GAPDH IgG concentrations in controls and cases. Values are shown as individual concentrations (controls, blue dots; cases, pink dots). Dashed line corresponds to the lower limit of quantification (LLOQ), 0.003 μg/mL. Statistical analysis was performed using an unpaired t test with Welch’s correction after logarithmic transformation of individual concentrations. Mean and 95% confidence intervals (CI) were calculated, and values were back transformed for graphical representation.

### Low concentrations of anti-GAPDH IgG are associated with higher susceptibility to bacteraemia

The quantification of anti-GAPDH IgG showed a significant reduction of these antibodies in individuals diagnosed with *E. coli* bacteraemia. To explore the influence of circulating anti-GAPDH IgG in the susceptibility to *E. coli* bacteraemia, we performed a logistic regression analysis in two steps. First, we evaluated the isolated effect of anti-GAPDH IgG on the diagnosis of bacteraemia. In the second step, although matching for age and sex had already been applied, we included these variables as potential confounders to minimize any residual bias.

As summarized in Table 3, the results from the unadjusted and adjusted models were very similar, showing consistent findings. In the adjusted model, each 1 μg/mL increase in anti-GAPDH IgG was associated with an OR of 0.18 (95% CI, 0.08-0.37). This reveals a strong inverse relationship between anti-GAPDH IgG concentration and *E. coli* bacteraemia diagnosis that corresponds to a reduction of 82% in the likelihood of bacteraemia per unit increase.

**Table 3.**
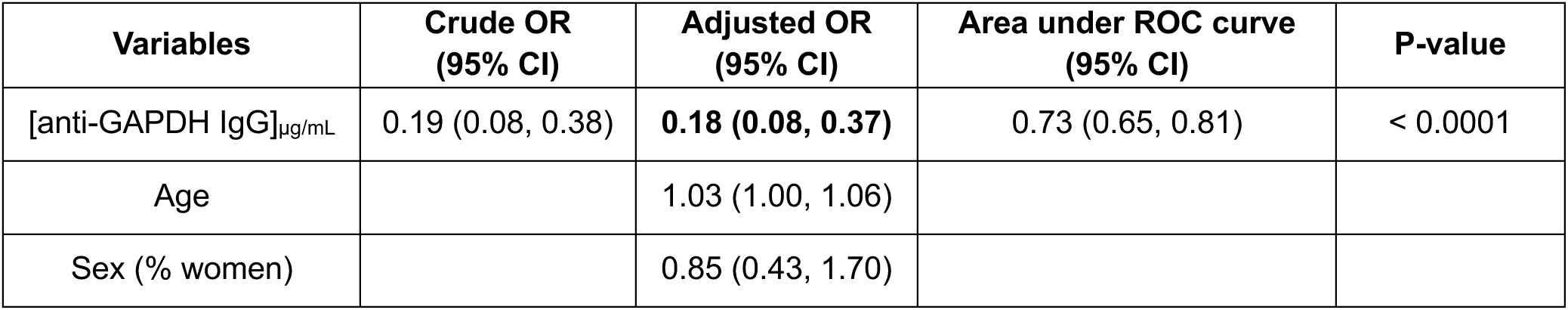
Association between circulating anti-GAPDH IgG and susceptibility to *E. coli* bacteraemia. Odds ratios (OR) and 95% confidence intervals (CI) were estimated using simple logistic regression (crude OR) and multiple logistic regression (adjusted OR). Predictive performance of the model was assessed using the area under the receiver operating characteristic (ROC) curve, with 95% CI and p-value reported.

| Variables | Crude OR<br>(95% CI) | Adjusted OR<br>(95% CI) | Area under ROC curve<br>(95% CI) | P-value |
| --- | --- | --- | --- | --- |
| [anti-GAPDH IgG]<sub>μg/mL</sub> | 0.19 (0.08, 0.38) | **0.18 (0.08, 0.37)** | 0.73 (0.65, 0.81) | < 0.0001 |
| Age |  | 1.03 (1.00, 1.06) |  |  |
| Sex (% women) |  | 0.85 (0.43, 1.70) |  |  |

The performance of the adjusted logistic regression model was evaluated using a ROC curve, which showed an area under de curve (AUC) of 0.73 (95% CI, 0.65-0.81; p-value < 0.0001) indicating good discriminative power of anti-GAPDH IgG in classifying individuals as cases or controls (Supplementary data Figure S2).

To further quantify the protective effect of anti-GAPDH IgG levels, we performed a logistic regression with categorised anti-GAPDH IgG concentrations, based on the distribution of concentration of anti-GAPDH IgG in the control group, as detailed in Methods section. As shown in Table 4, increasing anti-GAPDH IgG levels were associated with progressively lower odds of bacteraemia. Compared with the reference group (< 0.10 µg/mL), higher antibody levels were inversely associated with *E. coli* bacteraemia, reaching up to an 96% reduction in the likelihood of infection. These results demonstrate a clear dose–response relationship, with higher anti-GAPDH IgG concentrations conferring progressively greater protection against infection.

**Table 4.** Association between anti-GAPDH IgG and *E. coli* bacteraemia by concentration category. Category thresholds were defined according to the distribution of anti-GAPDH IgG concentrations in the control group based on quartiles: Q1, very low levels; Q2, low levels; Q3, high levels. Odds ratios (OR) and 95% confidence intervals (CI) in each category were calculated relative to the reference group (< 0.10 μg/mL). Relative OR reduction (1 − OR) is shown for each category.

| anti-GAPDH IgG |  | OR (95% CI) | Relative OR reduction |
| --- | --- | --- | --- |
| Categories | Conc. µg/mL |  |  |
| Reference | < 0.10 µg/mL (reference) | ---- |  |
| Very low levels | 0.10 to < 0.22 µg/mL | 0.21 (0.07, 0.59) | 0.79 |
| Low levels | 0.22 to < 0.38 µg/mL | 0.12 (0.04, 0.34) | 0.88 |
| High levels | 0.38 to < 0.72 µg/mL | 0.10 (0.03, 0.29) | 0.90 |
| Very high levels | ≥ 0.72 µg/mL | 0.04 (0.01, 0.15) | 0.96 |

### Estimated threshold of protection against *E. coli* bacteraemia

Bacterial GAPDH has been proposed as a promising vaccine target, with preclinical studies reporting successful outcomes from formulations designed to induce anti-GAPDH antibodies (27). Although the efficacy and immunogenicity of a vaccine targeting bacterial GAPDH can only be fully evaluated in clinical trials, the findings from this study allowed us to predict the impact of anti-GAPDH IgG in the protection from *E. coli* bacteraemia.

Using the logistic regression model evaluating the effect of anti-GAPDH IgG concentrations, we were able to predict probabilities of *E. coli* bacteraemia based on anti-GAPDH IgG concentrations. As observed in Figure 4, the progressive increase in anti-GAPDH IgG concentration promotes a reduction in the predicted probability of developing *E. coli* bacteraemia which supports the efficacy of anti-GAPDH IgG in the protection from infections caused by *E. coli*. We also calculated probabilities using the model adjusted for age and sex, fixing the values at the median age (70 years) and the most frequent sex (female), as shown in Supplementary Figure S3. The results were very similar to those obtained with the unadjusted model.

**Figure 4.**
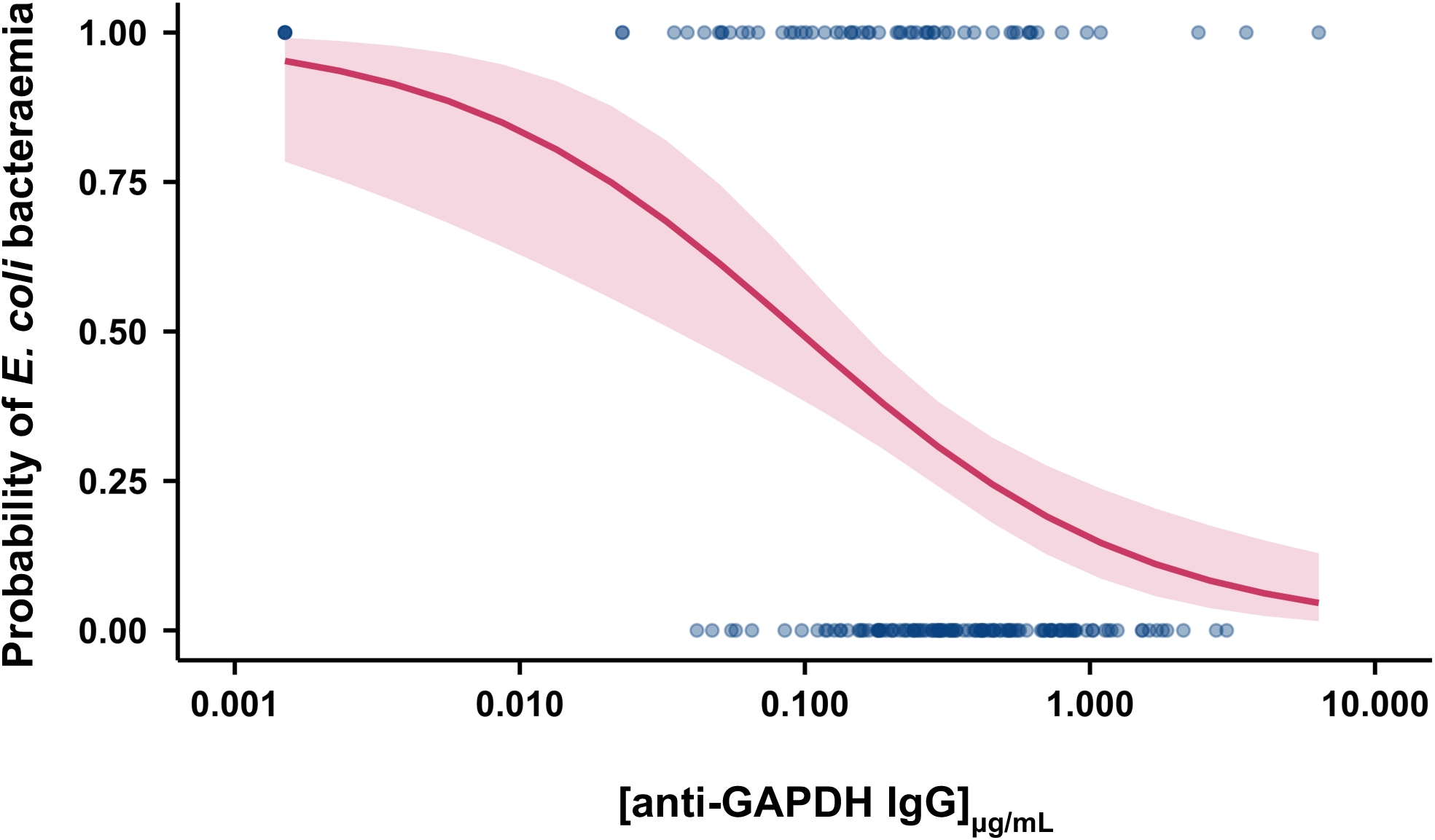
Predicted probability of a diagnosis of *E. coli* bacteraemia according to circulating anti-GAPDH IgG concentration. The solid red line represents the predicted probability of *E. coli* bacteraemia estimated using simple logistic regression, and the shaded area represents the 95% confidence interval.

In line with these findings and analysing the distribution of cases according to their anti-GAPDH IgG concentration, we were also able to estimate a protective threshold for anti-GAPDH IgG concentration. By analysing the distribution of cases according to their anti-GAPDH IgG concentration, we identified a concentration at which the number of infected individuals levelled off. As observed in Figure 5, we found that 95.2% of the cases had anti-GAPDH IgG concentrations equal or below 1.0 μg/mL, indicating a concentration threshold that appears to be associated with reduced susceptibility to *E. coli* bacteraemia.

**Figure 5.**
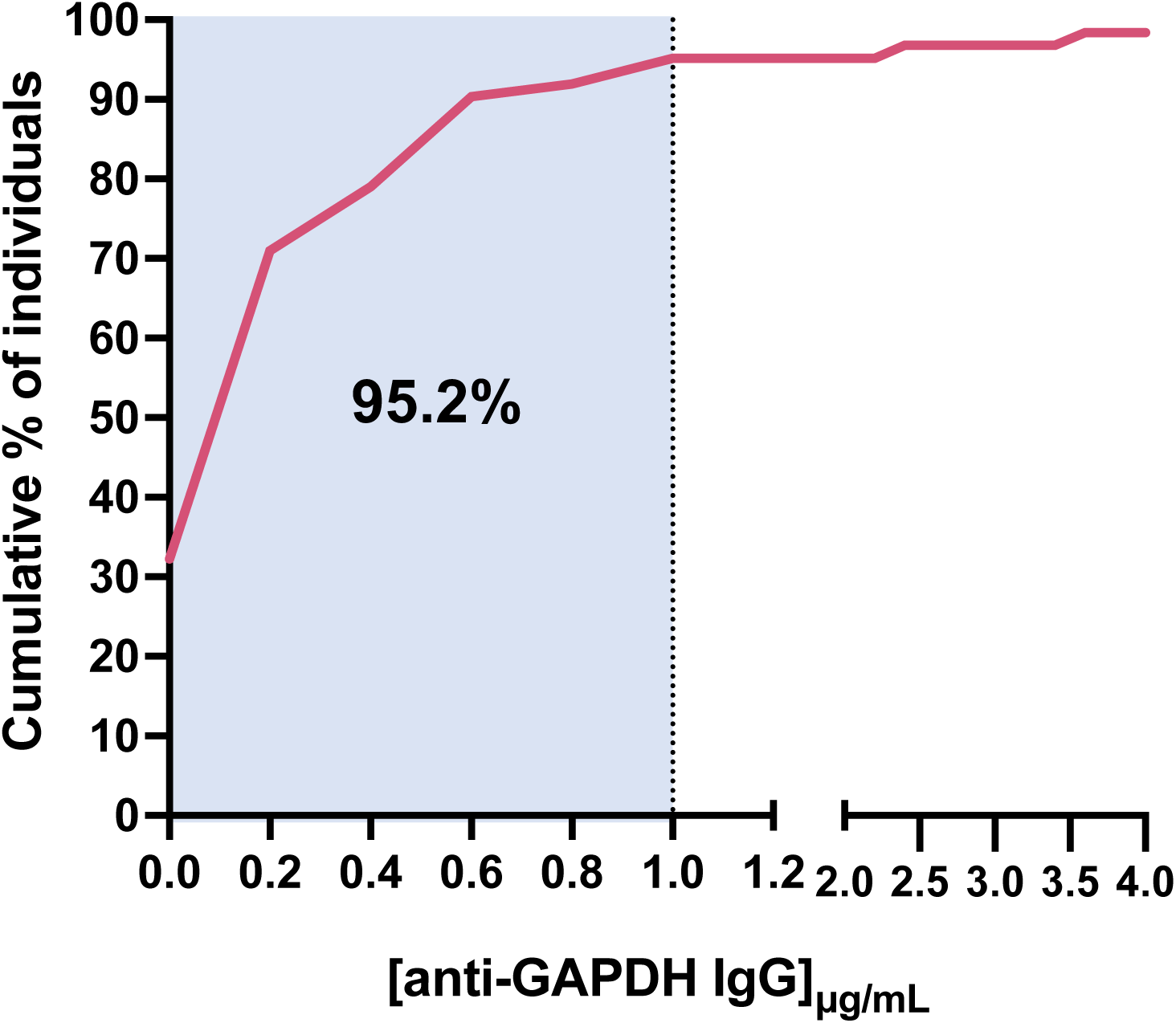
Cumulative distribution of circulating anti-GAPDH IgG in cases. The solid pink line represents the cumulative percentage of concentrations of anti-GAPDH IgG in individuals diagnosed with *E. coli* bacteraemia. Vertical dashed lines indicate the 95.2% of individuals whose anti-GAPDH IgG concentration is below 1.0 μg/mL.

### IL-10 and anti-GAPDH IgG as predictors of individual risk of *E. coli* bacteraemia

The results obtained at this stage showed us that individuals with *E. coli* bacteraemia generally exhibit detectable IL-10 in plasma and lower circulating anti-GAPDH IgG levels comparing with controls, and that the progressive increase in anti-GAPDH IgG levels reduces the probability of *E. coli* bacteraemia. Accordingly, we found a significant association between detection of IL-10 and low levels of anti-GAPDH IgG (Chi-squared test, p-value = 0.0028, Table 5), indicating that these variables are not independent and can be combined as a diagnostic marker.

**Table 5.** Association between low circulating anti-GAPDH IgG levels and the detection of plasma IL-10. Contingency table shows the number and percentage of individuals (n, %) with low or high anti-GAPDH IgG levels according to IL-10 detection (Not detected or Detected). Statistical significance was assessed using the Chi-squared test, and the p-value is reported.

|  |  | IL-10 |  | Chi-squared test |
| --- | --- | --- | --- | --- |
|  |  | Not detected | Detected |  |
| Levels of anti-GAPDH IgG | Low | 65 (60.2 %) | 43 (39.8 %) | 0.0028 |
|  | High | 63 (80.8 %) | 15 (19.2 %) |  |

We then fitted a multivariate logistic regression model including both IL-10 detection and low anti-GAPDH IgG levels, adjusted for age and sex. Since the study design already matched for these variables, results from the adjusted and unadjusted models were highly similar.

Therefore, we present the unadjusted model in Table 6 (adjusted analysis in Table S3, Supplementary data). The model confirmed the strong impact of IL-10 detection on infection likelihood (OR = 629, 95% CI, 139–5355). Low anti-GAPDH IgG levels were also associated with increased risk (OR = 4.26, 95% CI, 0.91–30.6), although this effect did not reach statistical significance in this categorical analysis. However, previous analyses evaluating anti-GAPDH IgG as a continuous variable demonstrated a significant inverse association with susceptibility to infection, further supporting its relevance as a potential diagnostic biomarker. The model exhibited excellent discriminative ability, with an AUC of 0.96 (95% CI, 0.93–0.99, p > 0.0001, Figure 6; Supplementary Figure S4 for the adjusted model), and predicted probabilities closely matched observed proportions, as shown by the calibration curve (Figure 7; Supplementary Figure S5 for the adjusted model).

**Figure 6.**
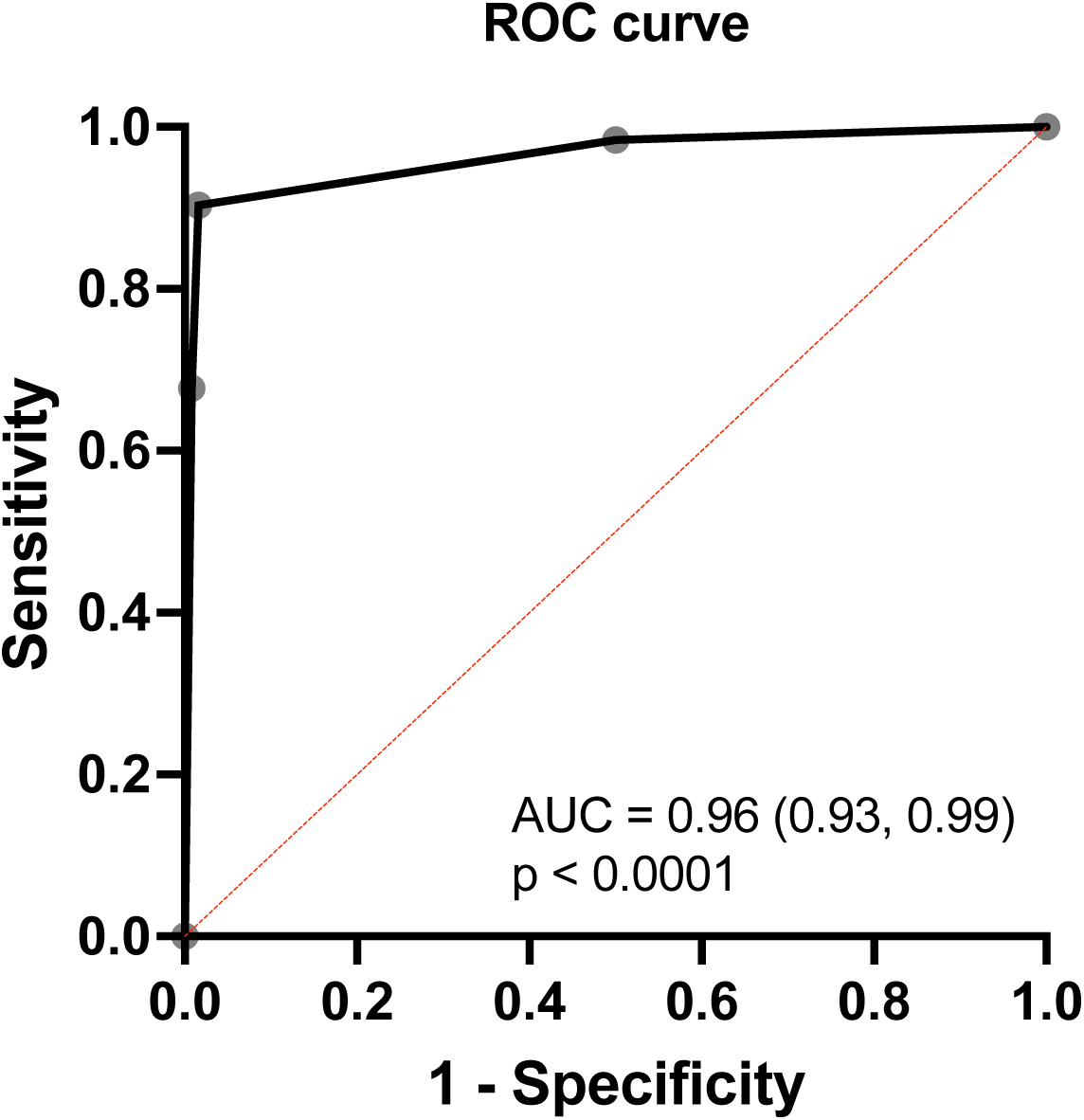
Receiver operating characteristic (ROC) curve for detection of IL-10 and low circulating anti-GAPDH IgG levels in *E. coli* bacteraemia. The curve was generated from the unadjusted model including both variables. Area under the curve (AUC) and p-value are shown within the figure.

**Figure 7.**
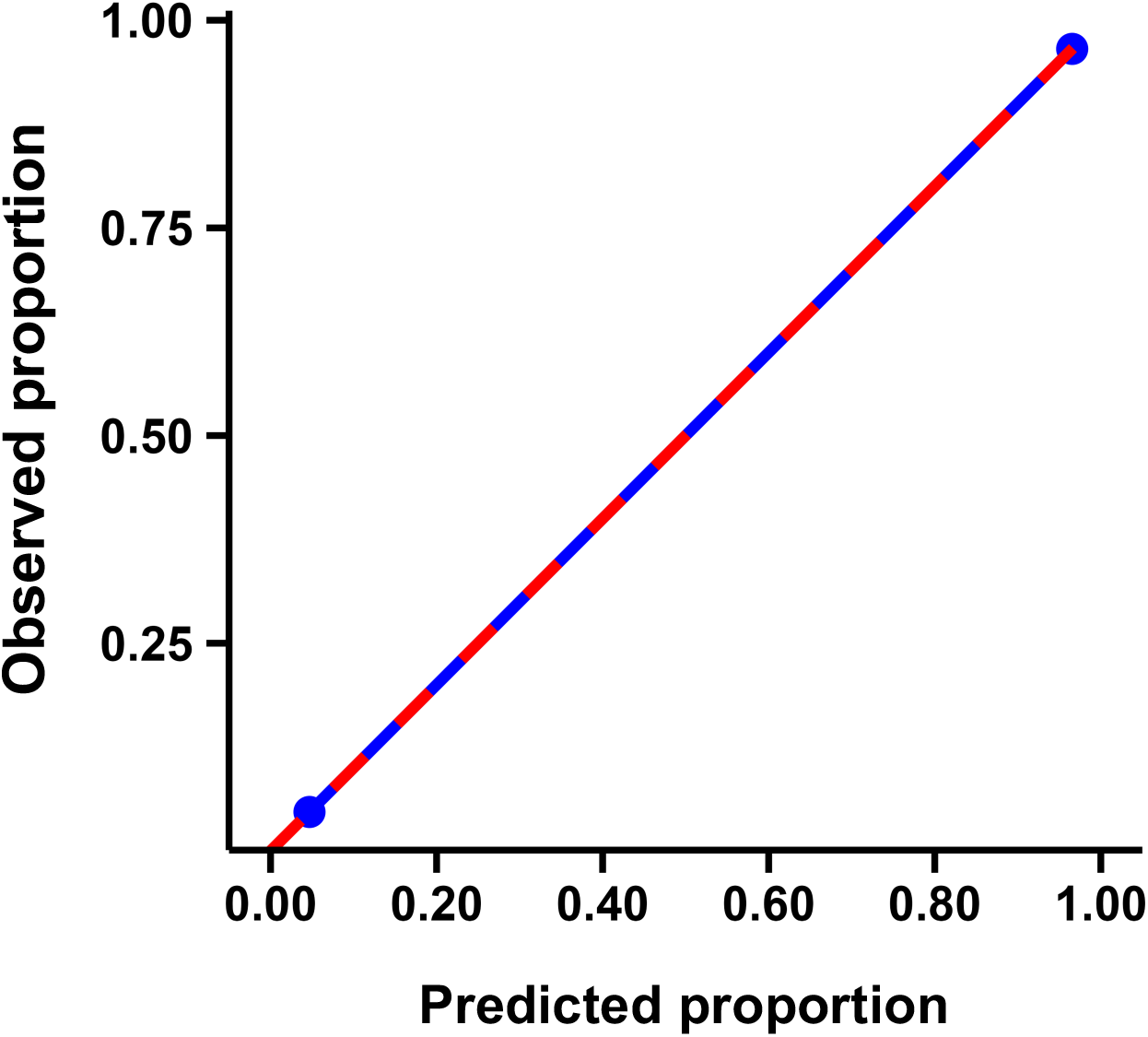
Calibration curve for detection of IL-10 and low circulating anti-GAPDH IgG levels in *E. coli* bacteraemia. The blue line represents the observed probability of *E. coli* bacteraemia within each quintile of predicted probabilities, based on the unadjusted model combining both variables. The red dashed line indicates perfect calibration.

**Table 6.**
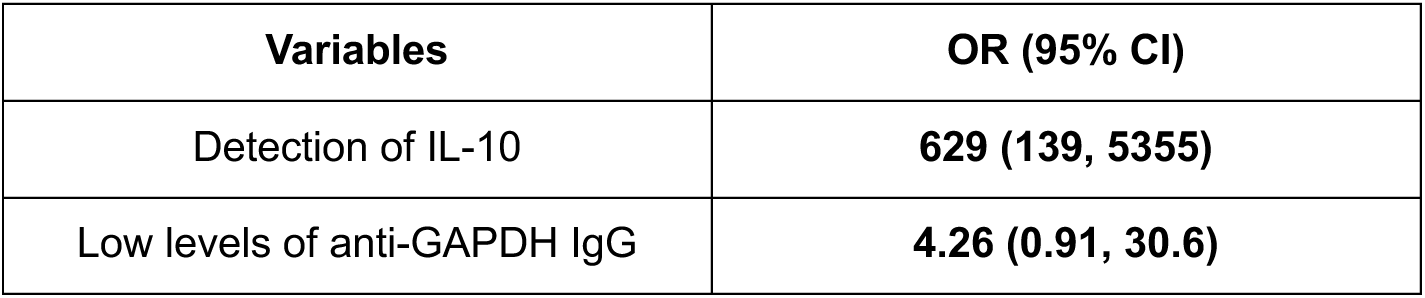
Association of detection of IL-10 and low circulating anti-GAPDH IgG levels with *E. coli* bacteraemia. Odds ratios (OR) and 95% confidence intervals (CI) are shown for each variable included in the unadjusted logistic regression model.

These results highlight that the combination of IL-10 and anti-GAPDH IgG provides robust discrimination between infected and non-infected individuals. Therefore, we used the unadjusted model to define four distinct biomarker profiles, representing all possible combinations of IL-10 detection (negative or positive) and anti-GAPDH IgG levels (low or high). As schematically represented in Figure 8A, this framework created a diagnostic matrix, providing a practical tool to estimate individual probabilities of *E. coli* bacteraemia based on IL-10 and anti-GAPDH IgG profiles, enabling early risk stratification and guiding timely therapeutic interventions.

**Figure 8.**
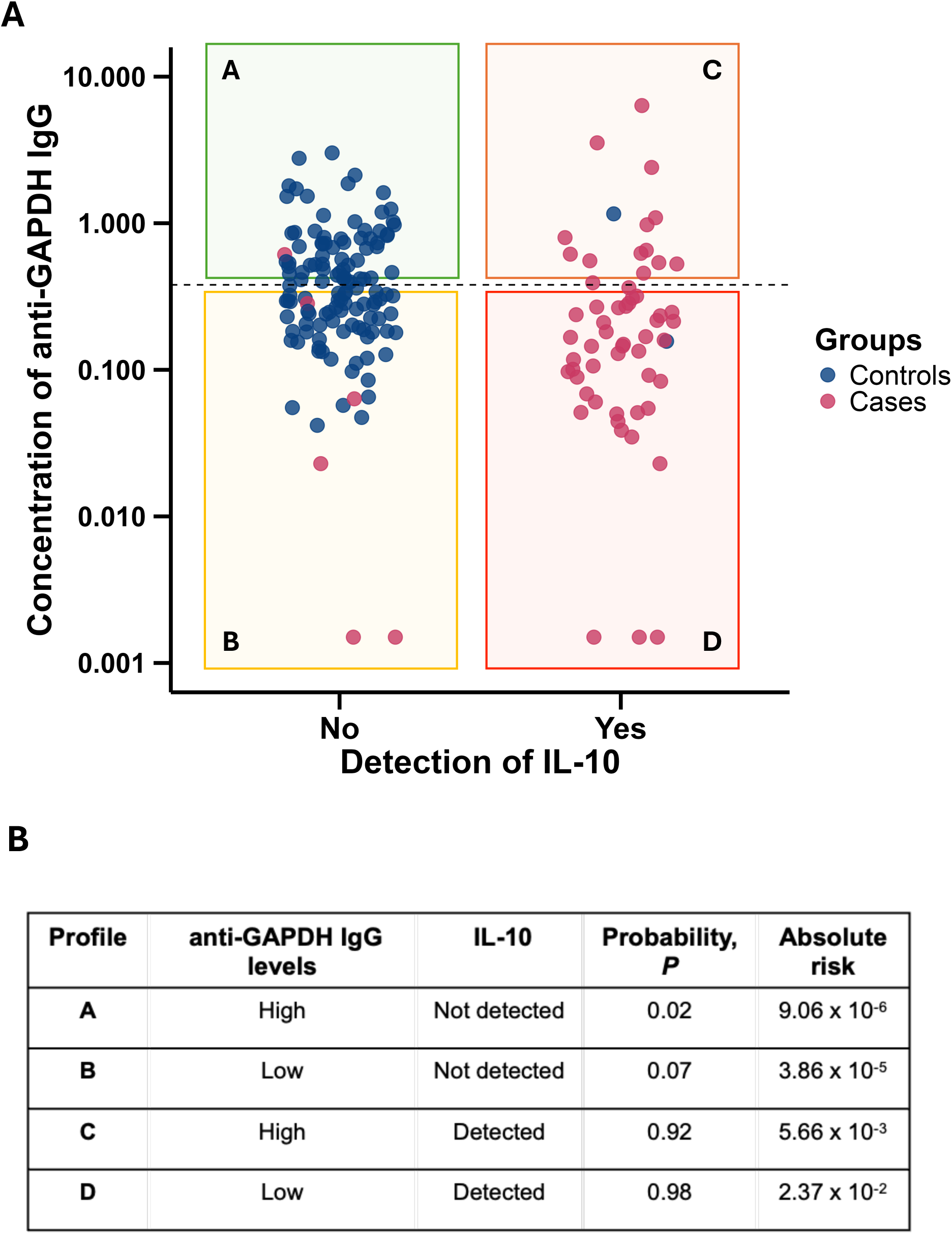
Predicted probability and absolute risk of *E. coli* bacteraemia based on IL-10 and anti-GAPDH IgG. **(A)** Individual concentrations of anti-GAPDH IgG in controls (blue dots) and cases (pink dots) distributed according to IL-10 detection. **(B)** Summary of the four profiles based on high or low anti-GAPDH IgG levels and IL-10 status (not detected or detected), showing the corresponding predicted probabilities calculated from the unadjusted logistic regression model and absolute risks, corrected for the actual population incidence of *E. coli* bacteraemia (48 cases per 100,000 individuals per year; Bonten et al., 2021 (30)).

To further contextualize these probabilities, we calibrated the model to the incidence rate of *E. coli* bacteraemia (48 cases per 10,000 person-years (30)), translating predicted probabilities into absolute risk estimates for each biomarker-defined profile. As shown in Figure 8B, both the predicted probability and absolute risk of infection increase with low levels of anti-GAPDH IgG and detection of IL-10. Individuals with low anti-GAPDH IgG and detectable IL-10 have the highest risk, while those lacking both markers have minimal risk of *E. coli* bacteraemia.

Detailed values for each biomarker-defined profile, including point estimates, medians, and 95% confidence intervals for both predicted probabilities and calibrated absolute risks, are presented in Supplementary Table S4.

## DISCUSSION

This observational study provides novel evidence about the impact of anti-GAPDH IgG in preventing human bacterial infections caused by *E. coli*. Our data shows that higher circulating levels of anti-GAPDH IgG are associated with a reduced susceptibility to *E. coli* bacteraemia. Over the past decades, several studies suggested a potential role of antibodies against GAPDH in conferring resistance to infections caused by bacteria or parasites (39–42). However, and to our knowledge, this is the first study that quantifies the impact of increased anti-GAPDH IgG levels on the probability of an individual to be diagnosed with *E. coli* bacteraemia.

Infections caused by *E. coli* represent a clinical threat for human health, partly due to the escalating AMR and the often difficult and delayed diagnosis of these infections. Additionally, the global ageing of the population increases vulnerability to such infections, and the development of effective vaccines against *E. coli* has faced considerable challenges, with recent clinical trials yielding disappointing results (9,10,30).

Different serological studies have consistently demonstrated that antibodies against structural epitopes of *E. coli* do not confer protection. Instead, patients with infections caused by *E. coli* or other *Enterobacteriaceae* typically exhibit higher antibody levels compared with non-infected individuals (13,14,16,19,20). For example, Brauner et al. (1989) reported increased titers against homologous LPS in patients with *E. coli* bacteraemia compared with healthy controls. Similarly, Nicolle et al. (1988) observed stronger antibody responses against both LPS and outer membrane proteins of the infecting strain in patients with UTI, with titers correlating positively with disease severity. More recently, Isobe et al. (2014) demonstrated that patients with haemolytic uremic syndrome caused by *E. coli* had elevated anti-O antigen antibody levels, particularly in those with more severe disease. Taken together, these findings indicate that antibody responses against structural epitopes are more useful for diagnosis or disease severity assessment than for effective protection against *E. coli*.

Moreover, this rise in antibody titers also fails to prevent recurrent infections (15,22). Instead, chronic exposure to the same pathogen promotes the expansion of antibody responses targeting structural epitopes, as illustrated by the increased levels of antibodies against distinct regions of LPS (lipid A and O-antigen) reported in patients with recurrent UTI (14,21). This accumulating serological evidence provides a rationale for the disappointing results of recent *E. coli* vaccine trials. Most of these candidates relied on structural antigens which, although well tolerated and immunogenic, induce antibody responses that are not protective.

Taken together, these observations strongly suggest that protection against *E. coli* bacteraemia requires targeting alternative, non-structural antigens. In the last years, we have been proposing that some bacteria – including *E. coli* - possess an extracellular or excreted form of GAPDH that acts as an immunosuppressor by stimulating an early and elevated production of IL-10 (23,25,26, unpublished observations). As others, we also found a clear and significant association between the diagnosis of infection and high levels of IL-10 (27–31). We were also able to identify circulating bacterial GAPDH in the blood of patients from whom pure *E. coli* colonies were isolated. These findings align with and reinforce the hypothesis suggesting the immunosuppressive role of extracellular GAPDH.

In this context, one plausible explanation for the lack of protection conferred by antibodies targeting surface structural antigens is that GAPDH-induced immunosuppression may impair the activation of an adequate pro-inflammatory response (28,43), thereby compromising the development of an effective immune defense against the bacteria. In this context, one plausible explanation for the lack of protection conferred by antibodies targeting surface structural antigens is that GAPDH-induced immunosuppression may impair the activation of an adequate pro-inflammatory response (28,43), thereby compromising the development of an effective immune defence against the bacteria. As a result of high levels of IL-10 at early time points of infection, even if antibodies targeting the bacterial surface are present, they are unable to effectively activate effector mechanisms of bacterial clearance such as opsonophagocytosis-mediated killing by competent cells. Therefore, these findings suggest that immunization strategies solely targeting surface antigens are insufficient and that it is essential to neutralize bacterial GAPDH to prevent the rapid and early induction of IL-10.

Notably, pre-clinical studies have already demonstrated the efficacy of a conjugated peptide-based vaccine, consisting of GAPDH-derived surface peptides from Group B *Streptococcus* and *S. aureus* conjugated with KLH (27). Lemos et al. demonstrated, using a murine model, that immunization with extracellular GAPDH peptides induces antibodies that trigger a protective inflammatory response, associated with inhibition of IL-10 production immediately after infection and improved survival of infected mice. Furthermore, in an *ex vivo* human cord blood infection model, they showed that the presence of anti-GAPDH IgG led to a sustained reduction in bacterial replication.

Although the efficacy of a vaccine inducing anti-GAPDH antibodies can only be fully assessed in clinical trials, our study provides the first estimation of their protective effect in humans. All analytical approaches applied in this study converge on the same conclusion: higher levels of anti-GAPDH IgG are associated with reduced susceptibility to *E. coli* bacteraemia, reinforcing the robustness of evidence and the biological relevance of anti-GAPDH IgG as a protective factor.

In addition to restoring the immune system’s ability to control infection, targeting GAPDH as a vaccine antigen also overcomes an important limitation posed by the diversity and distribution of serotypes, which is a challenge when selecting serotype-specific antigens for vaccination. As further confirmed by the high degree of conservation observed among the extracellular GAPDH proteins of the *E. coli* isolates analysed in this study, this is a highly conserved protein across different serotypes and even among distinct bacterial species (23,25,26). Such conservation strengthens the potential of extracellular bacterial GAPDH as a broadly protective antigen, capable of overcoming serotype replacement and contributing to the prevention of infections caused by a wide range of bacteria. Beyond its vaccine potential, extracellular bacterial GAPDH also holds promise as a diagnostic biomarker. Its early secretion during infection enables rapid detection, providing a targeted alternative to conventional culture-based diagnostics, which are time-consuming and often require empirical use of broad-spectrum antibiotics, a practice that contributes to the increasing burden of AMR (43).

In line with this diagnostic perspective, our study developed a combined biomarker integrating plasma anti-GAPDH IgG levels and IL-10 detection. Given the inverse relationship between low levels of anti-GAPDH IgG and higher probability of infection and the strong association between IL-10 presence and *E. coli* bacteraemia, we constructed a diagnostic matrix integrating these two parameters. This combined biomarker represents a promising tool to estimate individual risk of *E. coli* infection and accelerate diagnosis, potentially guiding earlier and more targeted therapeutic interventions.

## CONCLUSIONS

This study highlights bacterial GAPDH as a promising vaccine target, whose neutralization can restore the ability of immune system to control infections caused by *E. coli*. For the first time in humans, we quantify the protective effect of anti-GAPDH antibodies and estimate how increasing these antibodies can counteract GAPDH-induced immunosuppression, thereby allowing pre-existing antibodies against bacterial surface antigens to act effectively. Together, these findings support a combined antibody strategy as a powerful approach to prevent *E. coli* infections, addressing a critical and urgent challenge in the prevention of bacterial infections.

## AUTHOR CONTRIBUTION STATEMENT

AF performed all experiments and data analysis. CT contributed to sample preparation. CN contributed to performing ELISA anti-GAPDH IgG quantification. JBN contributed to study design and bibliographic review. PC was responsible for recombinant protein and antibody production and purification. HO performed the mass spectrometry-based proteomics. FL, PC, CT and HO interpreted and discussed the proteomic results. SBM and CIS were responsible for the selection and provision of control samples. CL discussed and reviewed the study design and statistical analysis. MV, PM and AF designed and discussed the experiments and results. MV and PM supervised the entire study. Draft manuscript preparation was performed by AF and PM. All authors reviewed the results and approved the final version of the manuscript for publication.

## DATA AVAILABILITY

All data supporting the findings of this study are available within the paper and its Supplementary Information. Requests for additional data should be directed to the corresponding author.

