## Supplementary data for "Low levels of circulating IgG against bacterial GAPDH and elevated IL-10 levels are associated with increased susceptibility to *Escherichia coli* bacteraemia"

**Supplementary Table S1. Pairwise identity and similarity between extracellular bacterial GAPDH sequences from *E. coli* isolates obtained from plasma of individuals diagnosed with *E. coli* bacteraemia.** Rows and columns correspond to individual isolates, and each cell shows the percentage of sequence identity and similarity between the pair of isolates. The reference sequence B7MMQ3 (UniProt accession number) is included for comparison.

| | S15EC | S70EC | S71EC | S73EC | S74EC | S75EC | S76EC | B7MMQ3 |
|---|---|---|---|---|---|---|---|---|
| | Identity (%) | | | | | | | |
| **S15EC** | | 99.7 | 99.7 | 99.1 | 98.8 | 99.7 | 97.9 | 99.4 |
| **S70EC** | 99.7 | | 100 | 99.4 | 99.1 | 100 | 98.2 | 99.7 |
| **S71EC** | 99.7 | 100 | | 99.4 | 99.1 | 100 | 98.2 | 99.7 |
| **S73EC** | 99.4 | 99.7 | 99.7 | | 99.1 | 99.4 | 97.9 | 99.7 |
| **S74EC** | 99.1 | 99.4 | 99.4 | 99.7 | | 99.1 | 98.8 | 98.8 |
| **S75EC** | 99.7 | 100 | 100 | 99.7 | 99.4 | | 98.2 | 99.7 |
| **S76EC** | 98.8 | 99.1 | 99.1 | 98.8 | 99.1 | 99.1 | | 97.9 |
| **B7MMQ3** | 99.7 | 100 | 100 | 99.7 | 99.4 | 100 | 99.1 | |
| | Similarity (%) | | | | | | | |

**Supplementary Table S2. Association between detection of IL-10 and the diagnosis of *E. coli* bacteraemia.** Contingency table shows the number and percentage of controls and cases with IL-10 detected or not detected (n, %). Statistical significance was assessed using Fisher's exact test, and the p-value is reported.

| | | Controls | Cases | Fisher's exact test |
|---|---|---|---|---|
| IL-10 | Not detected | 122 (98.4%) | 6 (9.68%) | <0.0001 |
| | Detected | 2 (1.61%) | 56 (90.3%) | |

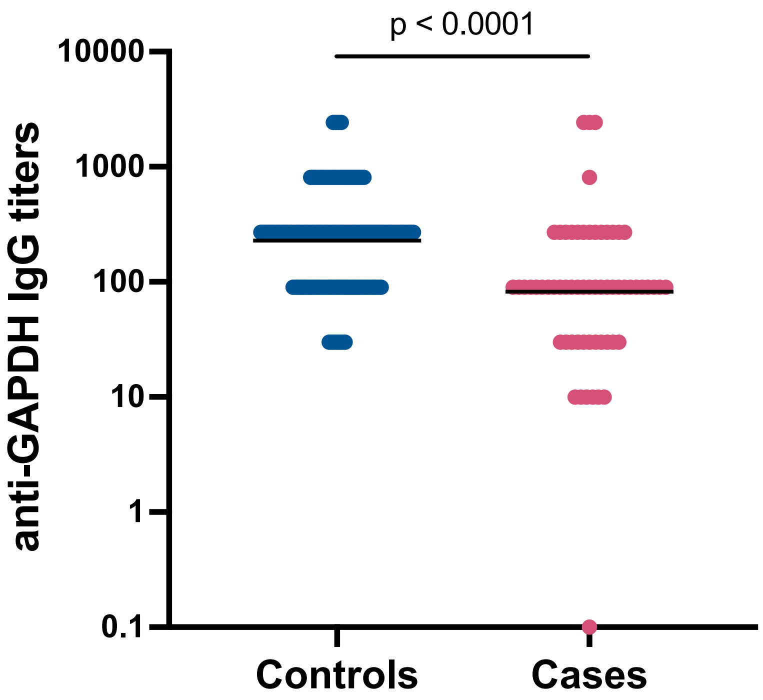


**Supplementary Figure S1. Circulating anti-GAPDH IgG titres in controls and cases diagnosed with *E. coli* bacteraemia.** Values are shown as individual titres, and horizontal bars indicate the geometric means. Statistical analysis was performed using an unpaired t test with Welch's correction after logarithmic transformation of individual titres. Mean and 95% confidence intervals (CI) were calculated and back-transformed for graphical representation.

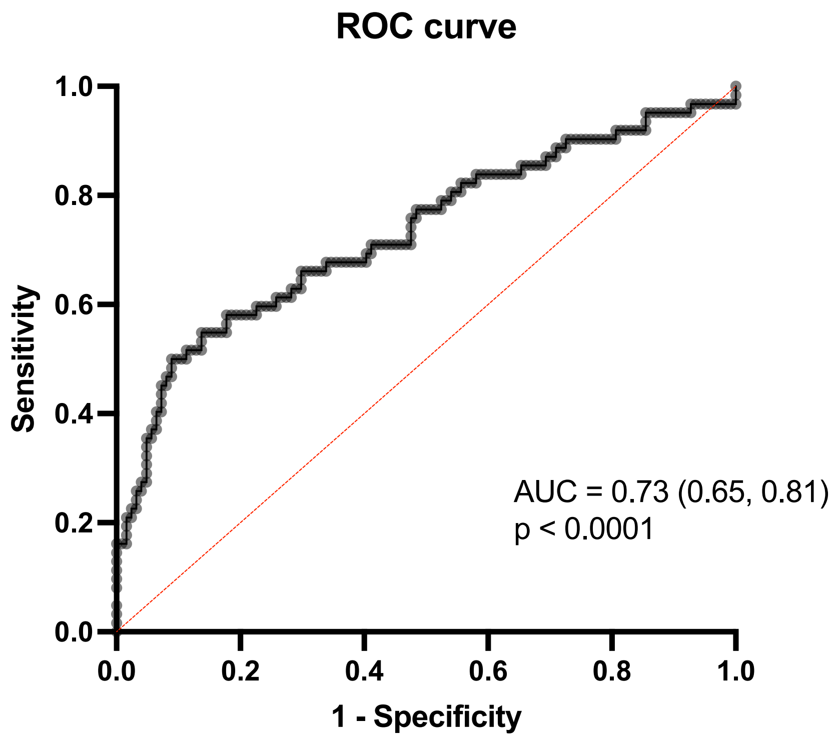


**Supplementary Figure S2. Receiver operating characteristic (ROC) curve for the logistic regression model predicting the diagnosis of *E. coli* bacteraemia based on circulating anti-GAPDH IgG concentration.** The logistic regression model was adjusted for age and sex. Area under the curve (AUC) and p-value are shown within the figure.

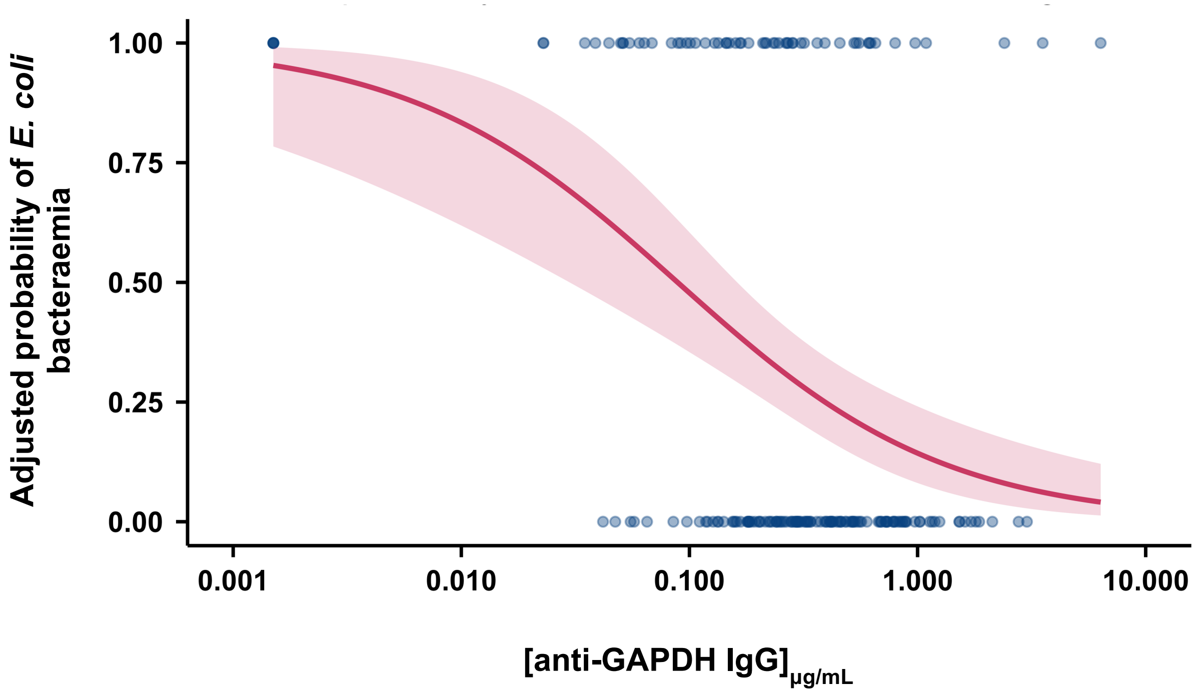


**Supplementary Figure S3. Predicted probability of a diagnosis of *E. coli* bacteraemia according to circulating anti-GAPDH IgG concentration.** Predicted probability was estimated using multiple logistic regression adjusted for age (fixed at the median, 70 years) and sex (woman, the most frequent). The solid red line represents the predicted probability based on the model, and the shaded area represents the 95% confidence interval (CI).

**Supplementar Table S3. Logistic regression model adjusted for age (years) and sex evaluating the combined association of detectable IL-10 levels and low circulating anti-GAPDH IgG levels with the diagnosis of *E. coli bacteraemia*.** Odds ratios (OR) and 95% confidence intervals (CI) are shown for each variable included in the model.

| Variables | Adjusted OR (95% CI) |
|---|---|
| Detection of IL-10 | **774 (152, 7962)** |
| Low levels of anti-GAPDH IgG | **4.12 (0.87, 29.9)** |
| Age | 0.99 (0.94, 1.05) |
| Sex (♀) | 2.08 (0.44, 16.7 ) |

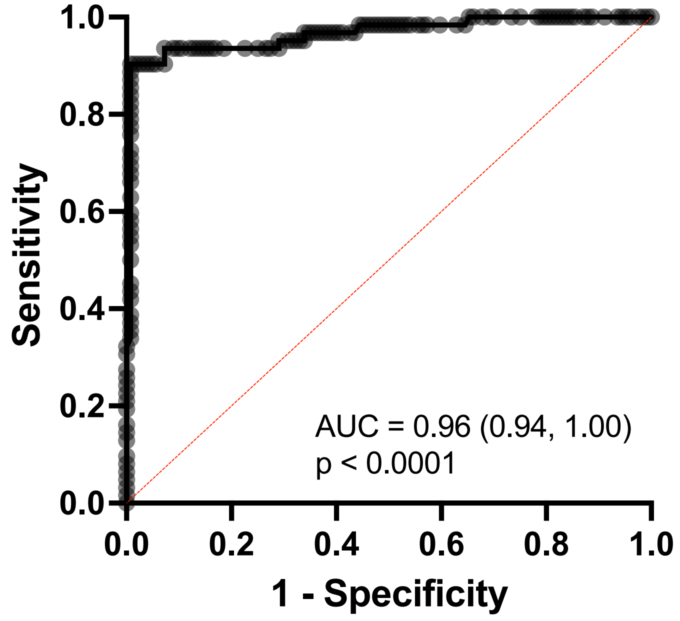


**Supplementary Figure S4. Receiver operating characteristic (ROC) curve for the logistic regression model adjusted for age (fixed at the median, 70 years) and sex (woman) evaluating the combined association of detectable IL-10 levels and low circulating anti-GAPDH IgG levels with the diagnosis of *E. coli* bacteraemia.** Area under the curve (AUC) and p-value are shown within the figure.

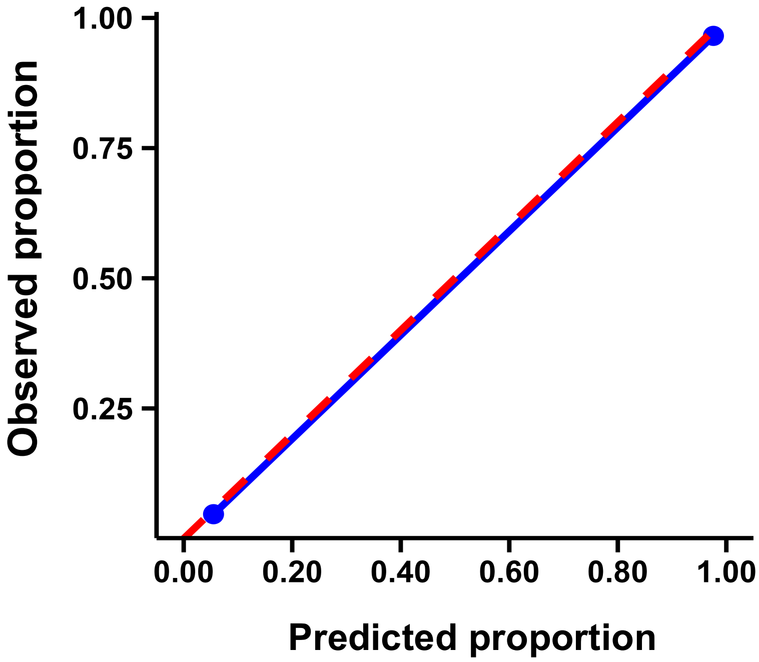


**Figure S5. Calibration curve for the logistic regression model adjusted for age (fixed at the median, 70 years) and sex (woman) evaluating the combined association of detectable IL-10 levels and low circulating anti-GAPDH IgG levels with the diagnosis of *E. coli* bacteraemia.** The blue line represents the observed probability of *E. coli* bacteraemia within each quintile of predicted probabilities. The red dashed line indicates perfect calibration.

**Supplementary Table S4. Point estimates, medians and 95% CI of predicted probabilities and absolute risks of *E. coli* bacteraemia according to IL-10 detection and low circulating anti-GAPDH IgG levels.** Each profile is defined by high or low levels of anti-GAPDH IgG and IL-10 detection status (not detected or detected). Predicted probabilities were calculated from the unadjusted logistic regression model, and calibrated absolute risks were derived using the population incidence of 48 cases per 100,000 individuals per year (Bonten et al., 2021 (30)). Values for each profile are presented as point estimate, median, and 95% confidence interval (CI) for both predicted probability and calibrated absolute risk.

| **Profile** | **Levels of anti-GAPDH IgG** | **IL-10** | **Predicted probability** | | | **Absolute risk** | | |
|---|---|---|---|---|---|---|---|---|
| | | | Point estimate | Median | 95% CI | Point estimate | Median | 95% CI |
| **A** | Low | Not detected | 0.018 | 0.019 | 0.004, 0.081 | $9.06 \times 10^{-6}$ | $9.15 \times 10^{-6}$ | $1.92 \times 10^{-6}$, $4.24 \times 10^{-5}$ |
| **B** | High | Not detected | 0.074 | 0.074 | 0.032, 0.164 | $3.86 \times 10^{-5}$ | $3.87 \times 10^{-5}$ | $1.57 \times 10^{-5}$, $9.46 \times 10^{-5}$ |
| **C** | Low | Detected | 0.922 | 0.922 | 0.704, 0.922 | $5.66 \times 10^{-3}$ | $5.66 \times 10^{-3}$ | $1.14 \times 10^{-3}$, $2.70 \times 10^{-2}$ |
| **D** | High | Detected | 0.981 | 0.981 | 0.899, 0.996 | $2.37 \times 10^{-2}$ | $2.39 \times 10^{-2}$ | $4.26 \times 10^{-3}$, $1.19 \times 10^{-1}$ |
